# Suppress, and not just flatten:Strategies for Rapid Suppression of COVID19 transmission in Small World Communities

**DOI:** 10.1101/2020.04.29.20085126

**Authors:** Chiranjib Bhattacharyya, V Vinay

**Affiliations:** Indian Institute of Science; Ati Motors and Chennai Mathematical Institute

## Abstract

Many countries have introduced Lockdowns to contain the COVID19 epidemic. Lockdowns, though an effective policy for containment, imposes a heavy cost on the economy as it enforces extreme social distancing measures on the whole population. The objective of this note is to study alternatives to Lockdown which are either more targeted or allows partial opening of the economy. Cities are often spatially organized into wards. We introduce Multi-lattice small world (**MLSW**) network as a model of a city where each ward is represented by a 2D lattice and each vertex in the latex represents an agent endowed with SEIR dynamics. Through simulation studies on **MLSW** we examine a variety of candidate suppression policies and find that restricting Lockdowns to infected wards can indeed out-perform global Lockdowns in both reducing the attack rate and also shortening the duration of the epidemic. Even policies such as partial opening of the economy, such as Two Day Work Week, can be competitive if augmented with extensive Contact Tracing.

## INTRODUCTION

The World Health organization(WHO) on March 12, 2020 declared COVID-19 outbreak as a pandemic^1^. The virus first emerged in China in late December [19], and since than it has invaded more than 150 countries posing an unprecedented threat to global public health. As of 19th April more than 2 million people have been infected globally and has resulted in more than 100 thousand deaths. The Director of WHO recently warned that *The worst is yet to come* (Press). Several countries have suffered massive loss of human lives, there are far greater number of countries where COVID19 has just arrived. The anticipated cumulative loss of lives in these countries could be apocalyptic and require urgent measures. There is now a growing view among scientists (Press) and many world leaders (UN press release) that **Rapid Suppression of COVID19 transmission** must be the way forward.

In the absence of any vaccine, governments across the globe are announcing various forms of moderate to extreme Social Distancing measures for containing this pandemic which ranges from Self-Isolation to imposing Lockdowns. Lockdown was first used in China for fighting the COVID19 pandemic. The effectiveness of such Lockdowns is still being evaluated [6], but there is now global consensus that it is probably the best tool for containment. Despite the success of Lockdown there are however growing concerns that Lockdown may have unintended consequences which could be devastating to the economy (Press). Unemployment is an inevitable outcome of economic downturn and it is no surprise that Lockdowns have resulted in significant job losses. For instance, in France half of private sector employees have been unemployed because of Coronavirus related Lockdown (Press). There is thus an urgent need for seeking effective alternatives to Lockdowns.

Targeted Social Distancing, such as isolating individuals who are in the contact network of an infected individual, fares better than Population wide measures in treating specialized diseases([10]). Usually the contact network of an individual is rarely known. However, in recent times, one can use Contact Tracing using GPS enabled smartphones to discover the underlying contact network. Preliminary results suggest that contact tracing and Case isolation tools can help in containing COVID19 if applied very early. However, if applied even slightly later than 3 weeks the epidemic spirals out of control[9, 11].

In this note we seek to exploit the network structure to obtain a comparative assessment of various Non-Pharmaceutical Policies (NPIs) as suitable alternatives to Lockdowns when applied in the early stages of the epidemic.

A summary of the main findings are as follows

1. Cities are spatially organized into Wards. Restricting the imposition of a Lockdown to infected wards seems to be the best suppression strategy and can be more efficient than Lockdowns.
2. Opening up the economy, like a 2 Day work-week, can be competitive with global lockdowns if there is extensive contact tracing.
3. Contact Tracing involves checking the state of the persons in the contact network of the suspect and then isolating them if neccessary. Our empirical results suggest that this mayn’t be as effective as Tracing the *Contacts and their Contacts(***TC2S**). The suggested **TC2S**strategy can substantially lower the attack rate, by more than 30%, over the prevalent Contact Tracing approach.
4. Apart from the study we also contribute **MLSW**, a flexible mathematical model, which can be used for studying policy interventions in a city. The relevant software would be made available soon.

**Disclaimer:** The aim of this report to make authorities aware of policy alternatives to Lockdown. The conclusions we draw are about the relative efficacy of various strategies and not about their absolute predictability. Also, our study does not consider ethical factors, issues related to civil liberties, and economic hardships of the proposed interventions.

## METHODS

Understanding the spread of Infectious Disease on a Network has emerged as an active area of research in Mathematical Epidemiology (see [12, 18] for a survey). Compared to traditional compartmental models [13] these models incorporate contact structure and are thus considered as more realistic for explaining epidemics. Empirical modeling of COVID19 outbreak suggests that Network based models such as Small world networks can be a better candidate to explain the spread of the disease [23]. Small world networks [22] was first introduced in epidemiology by [1] for understanding epidemic spread on networks. Since then there has been substantial interest in using Small World Networks in modeling specialized disease outbreaks [15, 16, 20]. To the best of our knowledge no study has attempted to understand the spread of COVID19 on small world networks. Previously studies of epidemic modeling on Small World Networks have used SEIR, as a compartmental model, on 1D and 2D lattice based models [4].

In this note we simulate a SEIR model on a Small World network to assess several NPIs as sustainable strategies for hindering the progress of COVID19 in a city. Small world networks posit a 1D or 2D lattice structure on the entire population with occasional long edges. Such networks will miss the point that the cities are not necessarily homogeneously structured but are often organized into spatial clusters, for example the city of London has 25 wards. The motivation for modeling the ward structure arises from the need for containment of disease at the ward level. Directing measures to target affected individuals should have the most effect in containment but unfortunately there mayn’t be enough technological support for executing such measures. Imposing a coarse measure such as Lockdown on an entire city is unsustainable as it would face compliance issues from the broader population. Ward level interventions may strike the right compromise in deriving sustainable containment policies. It is also to be noted that ward wise containment policies are already being considered in several Indian cities including Bengaluru (Press). Our aim is to compare such strategies to Lockdown.

**Fig. 1.**
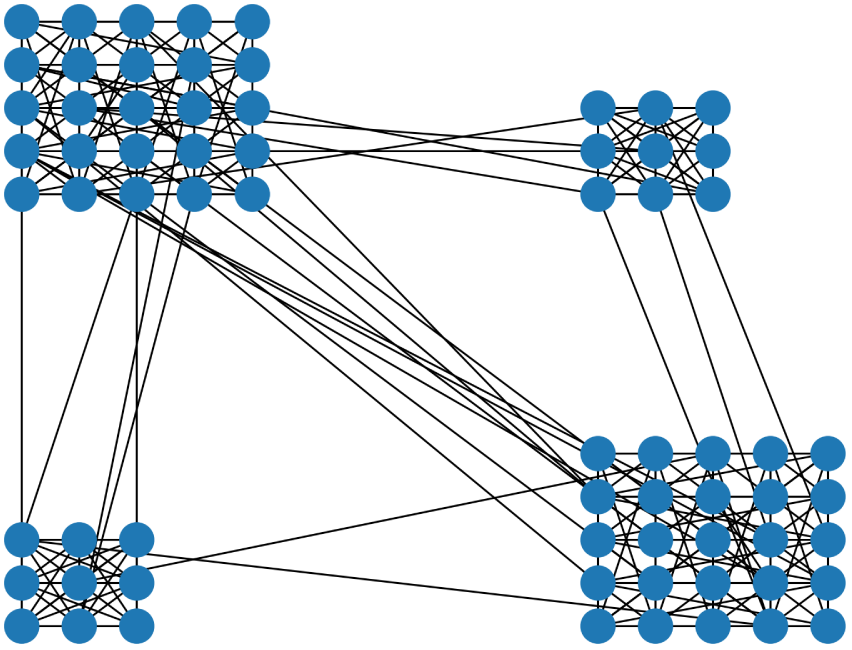
A multi lattice small world example with four wards. Each ward is a 2D torus.

### Urban Environments as Multi-Lattice Small worlds

We develop a small world model which involves modeling a ward as a 2D lattice. The city of Bengaluru is divided into 198 wards. Neighboring wards are naturally defined and they create edges in the resulting neighborhood graph of the city. Each ward is replaced by a population between 400 to 800 people in direct proportion to the population of each ward to simulate a city of roughly 10^5^ citizens.

We will now describe a small world model of a city consisting of wards. At each ward, the population is a regular 2D lattice with edges to all eight neighbors. This models local spatial interaction. The lattice is also wrapped around (it is a torus) so that it has no boundary. If two wards are neighbors, e vertex pairs are choosen at random across the two wards and an edge is inserted across each of the five pairs. These edges model interaction between adjacent wards. This is done for every pair of neighboring wards. In this way, the graph is created. What this graphs lacks are *l*ong edges. Inspired by the small world model, we rewire an edge in this graph with probability *p*_0_. We cycle through all the edges. Every edge (*u, v*), is rewired with probability *p*_0_. Here, we choose a new vertex *v′* randomly from the entire graph and the edge (u, v) is replaced by this edge (*u*, *v′*). These edges capture non-local contact interaction.

This procedure results in a graph which at one level contains neighborhood information of the city and at another, the small world characteristics due to the long edges. Since we have a collection of lattices in our graph (instead of one), we call it *M*ulti-Lattice Small World (**MLSW**). **MLSW** is parametrized by *{n*_1_, …, *n_W_*, **W**, **e**, *p*_0_} where **W** is the number of Lattices. A Lattice *w* ∊ {1,…, **W**} has *n_w_* vertices which reflects the population in the ward. Inter lattice edges are parametrized by **e** and finally *p*_0_ is the rewiring probability.

The city of Bengaluru when viewed as **MLSW** have *W* = 198, **e** = 5,400 ≤ *n_w_* ≤ 800, *p*_0_ = 0.1. The resulting graph has 116, 631 vertices and 472, 319 edges. This model can be enhanced in several ways by using detailed knowledge of the city such as transportation patterns to determine the long edges. Modeling Epidemics in Urban networks is not new (see [7] and citations therein). However, we are unaware of any work which tries to account for the ward structure. The proposed model, **MLSW**, is novel not only in epidemiology but also in the area of small world models. **MLSW** can be viewed as a special instance of *Random Spatial Networks* [2]. A detailed mathematical study comparing **MLSW** with such networks will be presented elsewhere. In this note we present a simulation study for assessing different NPIs as suppression policies.

### Disease progression and Transmission

In this section we develop an SEIR based state model of an stochastic Agent which resides in one of the vertices of **MLSW** and interacts with its neighbors. The parameters of the agent are adjusted to model the progression of the disease in an individual and the interaction between the agents are modeled to suit the transmission dynamics of COVID19.

### Disease Progression

We develop a state based model for Disease Progression in an Individual based on the following assumptions based on existing literature, also summarized in Table 1.

**Table 1.**
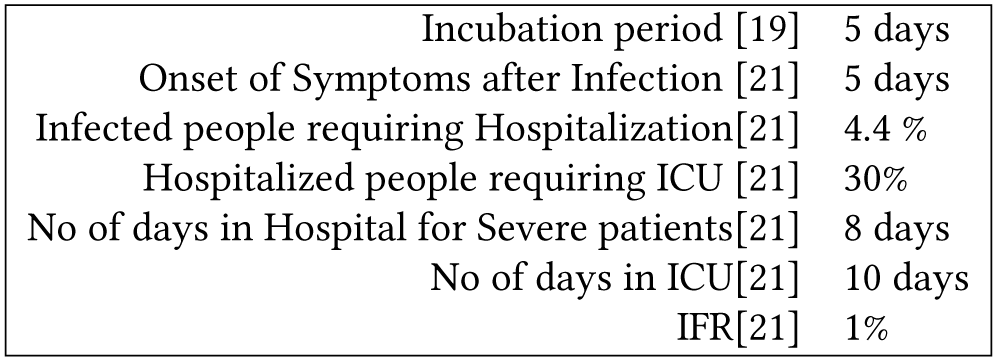
Assumptions on COVID 19 Disease progression

**Assumptions:** Incubation period of the virus is assumed to be 5 days. 4.4% of infected patients require hospitalization in 5 days after showing signs of infection. 30% of Hospitalized patients may require ICU admission or ventilator access. Half the ICU patients die.

**The state model:** Each vertex can be in one of seven states. All nodes start as susceptible S, and eventually end as either dead *D* or recovered *R*. In between a vertex will be either exposed *E* or infected *I*. We have three levels of infection, mild *IM*, severe *IS*, and critical *IC*. The entire state transition diagram including transition probabilities is defined in Figure 2. Based on the probabilites on the self loops, it is clear that the expected number of days to escape at nodes *E*, *IM*, *IS*, *IC* is 5, 5,8,10 respectively. These numbers are in tune with the number in [5].

**Fig. 2.**
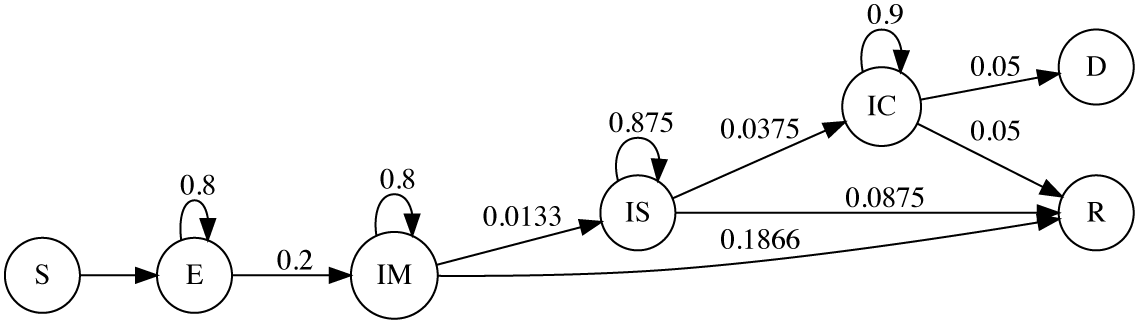
State Transiton Diagram

### Disease Transmission

To describe the spread of the disease we consider the following approach. When one end of an edge is infected (i.e., in state *IM, IS*, or *IS*), and the other end is susceptible *S*, with some probability, vertex at the other end transitions to *E*. This probability depends on the nature of infection and we will parametrize the probability of *IM* infecting a susceptible person to be *β_M_*. Similarly, we also introduce parameters *β_S_, β_C_* to measure probability of infection by a patient in state IS and IC respectively. We will calibrate *β = [β_M_, β_S_, β_C_*]^⊤^ to match the basic reproduction rate, *R*_0_, assumed to be 2.8 [14].

## SIMULATING THE PROGRESS OF COVID19 ON MLSW

The simulation begins by initializing the states of INDEX number of nodes to *IM* in **MLSW**. This state serves as introducing INDEX number of Index patients in the population.

A *day* in the Simulation consist of two steps, namely **Edge Sweep** and **Node Sweep**.

**Edge Sweep:** All the edges in **MLSW** are inspected in an arbitrary order. If edge *(u, v*) has one of its endpoints (say *v*) as susceptible S, and the other endpoint (*u*) as infected (there are three possibilities here: *IM*, *IS*, *IC*). Then the infection will spread to *v* with a probability that depends on the nature of the infection at *u*. Let *β* = [*β_M_*, *β_S_*, *β_C_*]^⊤^ be a vector of parameters where *β_M_*, *β_S_*, *β_C_* are probabilities of infecting a node with vertex *S* corresponding to states *IM*, *IS*, *IC* respectively. If the infection should spread, the new state of the node v would be exposure *E*.

**Vertex Sweep:** All nodes of **MLSW** are inspected in arbitrary order. Each node *u* is in one of seven states. Unless the states are in *R* or *D* where nothing happens as these are final states, in all other case the node transitions to a new state or stays put according to the probabilities assigned in 2. This mimics the progression of the disease in an individual.

The simulation runs through several *days* till there are no infected patients in the network, i.e. all states are in any one of *R*, *S*, *D* states.

## MODEL CALIBRATION

The **Edge Sweep** requires *β*. The state transition has 11 parameters as can be read from Figure 2. And then there is the rewiring probability *p*_0_. Each of these sets of parameters play a unique role.

The rewiring probability *p*_0_ determines how fast the disease can spread in the network. If the long edges are missing, we are left with lattices connected to their neighbors and this takes infection a long time to propagate. Long edges shorten this time. Even mild values of *p*_0_ show rapid ability to mix. Large *p*_0_ essentially results in a random graph.

The **Edge Sweep** parameters *β_M_*, *β_S_*, *β_C_* determine how fast the infection actually propagates on **MLSW**. The state transition matrix determines how long a person is active to infect. It also determines the death rate amongst the people who do get infected.

The lattice we have chosen has 8 neighbors corresponding to the eight immediate vertices around a fixed vertex on a 2D grid. Let us fix a vertex. It has typically 8 neighbors on an average (the specific degree will change a bit depending on the rewiring). Suppose the vertex is in state *IM*. Then the probability of propagating the infection to a fixed neighbor is *β_M_*. If the number of susceptible neighbors is d then the expected number of infections is *dβ_M_*. If the probability of exiting *IM* is γ*_M_* (this value can be read from the state transition diagram), the number of days it takes to exit the state is 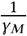. This means the total number of neighbors infected in expectation is 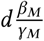. The process repeats itself for severe and critical infections but the dominant term is 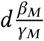. We now match this to the assumed value of *R*_0_ = 2.8 to obtain *β*_0_*_M_* = 0.07. In absence of any other data on hospital infections etc we set the value of *β*_0_*_S_* = 0.02 and *β*_0_*_C_* = 0.001. Hence, we will assume that *β* = *β*_0_ = [*β*_0_*_M_*, *β*_0_*_S_*, *β*_0_*_C_*]^⊤^.

An important parameter to match is the Infection Fatality Rate (IFR) rate. The IFR rate is the probability of transitioning from *E* to *D* in the state transition diagram. We match IFR rate, the percentage of total infected patients match, along with active infected patients peaking around 75 days, with those of [5]. In all our simulation INDEX is set to 5.

## INTERVENTION POLICIES FOR RAPID SUPPRESSION

We are seeking policy alternatives to Lockdown which can *rapidly suppress* the COVID19 pandemic. Lockdown measures, though extremely effective, involve extreme social distancing measures for the entire population hence making them unsustainable for long duration. Policy alternatives to Lockdowns should therefore seek more targeted Social distancing, or aim to limit the amount of time for continuous lockdown. Keeping this in mind we explore two kinds of strategies. The first kind of strategy, called Targeted Interventions, involve either *Contact Tracing and Isolation* or *Hotspot specific interventions*. The second kind of strategy involves limiting the severity of Lockdowns by various means such as opening the economy for a few days in the week, or relaxing the lockdown. A list of few envisaged policies are mentioned in Table 2.

**Table 2.** List of Interventions. TI stands for Targeted Interventions, RD stands for Relaxed LockDowns

| Policy | Type | Description |
| --- | --- | --- |
| **LD** |  | Lockdown |
| **LD(mild)** | RD | Mild Lockdown |
| **FDLD** | RD | 40 day lockdown |
| **TDWW+LD** | RD | Two day week followed by a strict lockdown |
| **TC2S** | TI | Tracing and sealing of neighbours and their neighbours of Infected patients |
| **WSO** | TI | Ward level sealing and opening with different relaxation factors |

### Metrics for Evaluating Suppression policies

Any suppression policy needs to be effective and sustainable. The *effectiveness* of any policy would depend on its ability to contain the infection to a small fraction of the population. Often, such measures could be extreme in nature and thus *sustainability* depends on the duration of the time it is enforced, shorter the better. To evaluate both these aspects one can consider the following metrics for evaluating a policy A.

- The length of the epidemic,denoted by *T_epi_(A)*, is defined as the duration of the epidemic starting with few infected patients and ending when there are no infected people in the population. For sustainability purposes *T_epi_(A)* should be low.
- Attack rate, measured as follows

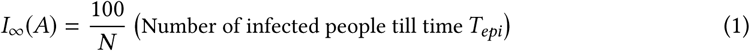 Since infected people either recover or die, *I*_∞_*(A) = R*_∞_*(A)* + *D*_∞_(*A*), where

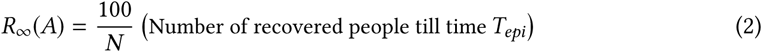

and *D*_∞_*(A)* denotes the percentage of Deaths due to policy *A*. A good suppression policy should ensure that *I*_∞_ is low which implies that both *D*_∞_ and *R*_∞_ are low.

From these observation one can draw the insight that a policy would gain acceptability if it is as effective as a lockdown but it is enforced for a shorter length of time than Lockdown. We encapsulate this insight via *preference score*, defined as follows.

*The preference score of a policy* A *with respect to a base policy* A_0_ *is defined as*

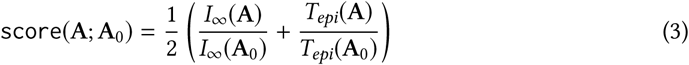

This score can be used to evaluate preference of a policy over the base policy A_0_. A policy A would be preferable to A_0_ if score(A; A_0_) is less than 1.

## DESCRIPTION OF INTERVENTION POLICIES

Using the designed metrics we will the study the acceptability of the following policies, also summarized in Table 2.

**Lockdown(LD)**: Lockdowns have been enforced in different ways across the globe. For example in UK (Press) you are allowed to go out for a walk or exercise once a day, while in India this is not allowed. There is also compliance To model this varied degree of Lockdown we introduce a parameter called *relaxation factor, η*, which is allowed to vary between [0, 1]. This parameter modulates the transmission parameter to obtain an effective transmission rate of *β =* η*β*_0_, The value of η = 0 corresponds to perfect lockdown and *η* = 1 represents the other extreme with no lockdown. Perfect lockdown corresponds to the entire population being quarantined. As practiced in many places **LD** is enforced by shutting down all but essential services. This should definitely lower considerably and getting such estimates would be difficult. One can assume that Lockdown will correspond to a small *η*. A small *η* corresponds to lowering of the reproduction rate in the network. (We hesitate to use *R*_0_ here because reproduction rate has meaning at any instant of time in a network.) In the absence of clear guidelines, in this note we will define **LD** as any measure which can achieve *η* ≤ 0.5. It would be useful to know how long should such a lockdown need to continue.

**Fixed Duration Lockdown(FDLD):** As an alternative to **LD** one can consider a fixed duration Lockdown (**FDLD**). In India, the Government announced a lockdown for 21 days on March 23rd and then extended it for another 20 days. Keeping this in mind we simulated a **FDLD** of 40 days and applied it after 20 days of onset of the pandemic.

**Tracing the Contacts and their Contacts and Sealing (TC2S):** Contact Tracing and Case Isolation is an important tool in the fight against this epidemic. It is long argued that Targeted Social Distancing may be more effective than social distancing measures imposed on the whole population [10]. Contact Tracing usually involves checking for infections in all persons who have come in the contact of an infected person. Based on empirical results (see Figure 4) we propose a two level strategy-not only trace the contacted persons of the patient, but also trace their contacts as well. We call this policy *Tracing the Contacts and their Contacts* (**TC2S**) and implement it as follows. The trigger is when a node *u* becomes severe *(IS*) or critical *(IC*). In this case, we look at all its neighbours *v* and seal it if it is state *IM*. We then look at the neighbours of v and then seal all their neighbours who is in state *IM*.as well. Finding the neighbours of *u* and their neighbours are part of the tracing mechanism. Sealing means the node will not have any connections with its neighbours and it could be implemented by either self-isolation or being admitted to Quarantine facilities. Here is a reason for doing this two level tracing. The node *u* must have got infected by one of its neighbours *u_p_* (who may have recovered since), who could have infected other neighbours as well. We cannot reach these neighbours by merely sealing the neighbours of *u*. Hence the two level tracing and sealing.

For such a scheme to be viable it is important that the number of people who are traced and isolated are small. We introduce a percentage measure of efficiency for **TC2S** as follows

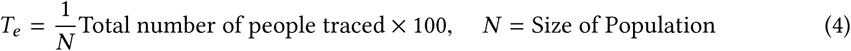

Very few countries have the ability to implement **TC2S** at the whole population scale and it is reasonable to assume that most countries can do it only when *T_e_* is a small fraction of the population.

**Ward Seal and Open(WSO):** In **LD** an entire city will be under lockdown. Instead of shutting the entire city one can enforce lockdowns in Infection Hotspots, in case of Bengaluru it could be wards with infected patients. Such strategies are already being considered (Press).

A ward can be isolated by removing all edges to other wards. In our case, we merely change *β_M_* by an η factor to account for account for delivery boys, health officials, and stray travel to groceries etc. The difference to the global lockdown is that the *β_M_* is lowered only for those nodes in wards that are in lockdown.

The opening and closing of ward are guided by a *low-water mark* and a *high-water mark*, denoted **WSO***(low, high)*. It will be assumed that once the infections in a ward cross the high-water mark the epidemic surveillance triggers are activated and the ward is declared as a Hotspot which requires intervention. The ward will be sealed that is lockdown will be imposed till such time that the infections subside to a low-water mark which signifies tolerable level. To measure the economic efficiency of **WSO** we introduce

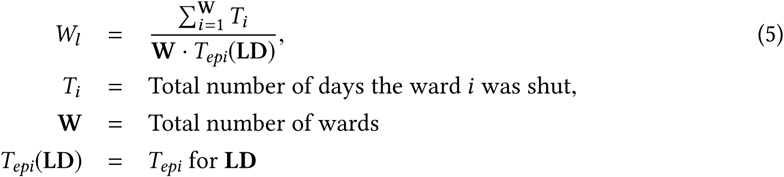

To explain the motivation for W_;_ we consider the following. Assume that loss to the economy incurred by shutting down one ward for 1 day is *c* units. The loss incurred by **WSO**is then 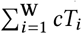. The loss incurred by **LD**policy is *cT_epi_***W**. Thus *W_l_* measures the relative loss to the economy compared to **LD**.

Thus any ward specific policy requiring the wards to be shut down should be considered practical only if *W_l_* is low. For our implementation we have chosen *low =* 0, and *high =* 3. For values of *low* other than 0 the system becomes unstable. Ideally one should aim for the highest value of the upper limit to minimize the duration of the lockdown.The chosen number 3 seems to be the best possible.

**Two day week followed by a Lockdown(TDWW+LD):** In spirit of **WSO** one can withdraw the lockdown for some amount of time and re-impose it after sometime. It is clear that with the infection doubling every 5 days[8], if left unchecked, it maybe risky to withdraw the lockdown for a whole week. Assuming that first symptoms will occur after 5 days, one could potentially allow a 2 day work-week. The policy intervention is to impose a lockdown for 5 days and open the economy for 2 days.

Other variations of the policies such as one Lockdowns on alternate weeks or three day week don’t seem to work as well as the two day week. So we don’t consider these alternatives here.

**Do Nothing(DN):** This is the base case where no interventions are done and the population achieves immunity after getting infected by the virus.

**Remarks:** We want to re-emphasis that we have taken a uniform approach to various stages of lockdown using a single factor *η*. For example, in a global or a ward level lockdown, it is natural to expect all long edges emanating from the ward to be ineffective. Instead, we retain the edges but dampen the probability of infection propagating to account for the delivery boys, the local police, the health workers etc who still move through the ward.

## RESULTS

In this section we present results of simulation studies of various policies mentioned in Table 2 on Small world network. One run of the algorithm starts with 5 index patients and iterates till there are no infected patients in the population. In Table 3 we report the median over 11 runs of relevant parameters namely projected number of deaths per million, *R_∞_*, and *T_epi_*. We also report *W_l_* and *T_e_* wherever applicable.

**Table 3.** *R_∞_*, *T_e_, W_l_* are reported in percentages(see (2),(4), and (5) respectively). *T_epi_* is the length of epidemic measured in days. All numbers reported are median values of 11 runs. Green colored cells indicate that the value in the cell is *acceptable*. Yellow colored cells indicate that the value *can be considered* while red colored cells indicate *unacceptable* values. For more discussion on the *acceptable, unacceptable, can be considered* see results. The policy acronyms are mentioned in Table 2

| | $\eta$ | Deaths<br>per million | $R_\infty$<br>% | $T_{epi}$<br>(days) | $T_e$<br>% | $W_l$ |
| --- | --- | --- | --- | --- | --- | --- |
| **DN** | 1 | 8702 | 84.96 | 178 |  |  |
| **FDLD** | 0.4 | 8479 | 84.88 | 232 |  |  |
| **TC1S** | 1 | 7528 | 74.77 | 209 | 11 |  |
| **TC2S** | 1 | 5642 | 55.63 | 303 | 15.3 |  |
| **WSO(0,3)(mild)** | 0.6 | 2992 | 29.97 | 480 |  | 3.78 |
| **LD(mild)** | 0.6 | 2675 | 25.75 | 557 |  |  |
| **TDWW+LD** | 0.4 | 1354 | 12.55 | 675 |  |  |
| **LD(mild)+TC2S** | 0.6 | 103 | 1.09 | 184 | 0.15 |  |
| **WSO(0,3)(mild)+TC2S** | 0.6 | 171 | 1.05 | 274 | 0.2 | .43 |
| **LD** | 0.4 | 51 | 0.52 | 115 |  |  |
| **TDWW+LD+TC2S** | 0.4 | 34 | 0.35 | 107 | 0.05 |  |
| **LD+TC2S** | 0.4 | 43 | 0.35 | 83 | 0.07 |  |
| **WSO(0,3)** | 0.4 | 18 | 0.33 | 124 |  | 0.18 |
| **WSO(0,3)+TC2S** | 0.4 | 18 | 0.09 | 65 | 0.02 | 0.04 |

For the sake of comparison between policies we have color-coded the values in the table. Red indicates *unacceptable*, Green indicates *acceptable* and *Yellow* indicates *can be considered*. We assume that a city’s Healthcare system can deal with patients comfortably if the percentage of infected population is less than 5%(*I*_∞_ is less than 5%). In case of epidemic outbreaks it can be pushed to higher values for a short amount of time say 15%. To assess the suitability of the duration of the policy, *T_epi_*, one could compare it with *T_epi_* for **LD**. From our simulations it seems that a strict Lockdown may take around 4 months(median value) to make the population Infection free. We rounded all *T_epi_* estimates to nearest months, and decide that all values less than 4 months are acceptable, as it is more efficient than **LD**. If *T_epi_* is more than double the duration during Lockdown, i.e. more than 8 months, we can consider it as *unacceptable*. Values in between can be deemedas *can be considered*. To assess the suitability of *T_e_* we assume that only a small fraction of the population can be contact traced and isolated. We set this limit to less than 0.5%. Any number more than 1% would be unacceptable. For *W_l_* one can argue that a policy should not be as expensive as **LD**. Any acceptable value must be necessarily less than 1. As lockdowns are expensive, one can argue that any policy should incur a loss which is a small fraction of the loss incurred by **LD**. We set the threshold for *W_l_* to be 0.1 for being acceptable. Any value between 0.1 and 1 *can be considered*. A value more than 1 is not acceptable. These thresholds are only notionally set to enable comparison. They can be set more precisely by using information from Health Departments from respective cities.

**Do Nothing, the base case:** Figure 3 shows the number of infections with time for **DN** when no interventions are applied. The peak occurs at about 67 days on an average. The number of deaths are 8702 per million and around 85% of the people have been infected and recovered. These numbers are in line with [5] and serves as a sanity check. The longevity of the virus seems to be roughly six months. The transmission parameters are unmodified with η = 1. Our numbers are slightly higher than that of [5] as we allow for Infections to be propagated by severely and critically ill patients as well.

**Fig. 3.**
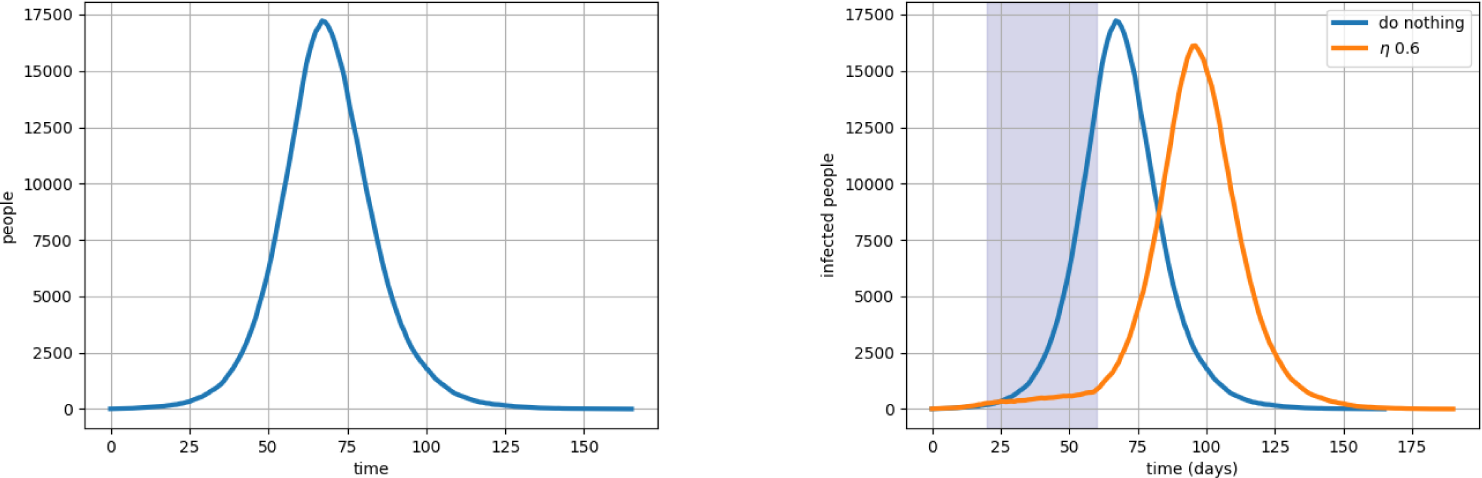
**DN** vs**FDLD**: On the left is the infection curve with no interventions. On the right is a forty day lockdown.

**Tracing and Sealing:** As can be see from Figure 4, **TC2S** as a stand alone strategy brings down the total infected population to less thab 2/3 of **DN** resulting in significant reduction of deaths per million. It also prolongs the life of the epidemic to a little less than a year. Fatality rate at 5650 deaths/million, though lower than **DN**, but still is unacceptably high. Also, it is to be noted that roughly 15% population needed to be sealed. To compare, the **TC1S** numbers are these: 7528 deaths/million while sealing 11% of the population. This is a 33% increase in death/million without a dramatic decrease in the percentage of population sealed. Hence we discard **TC1S** in favour of **TC2S**.

**Fig. 4.**
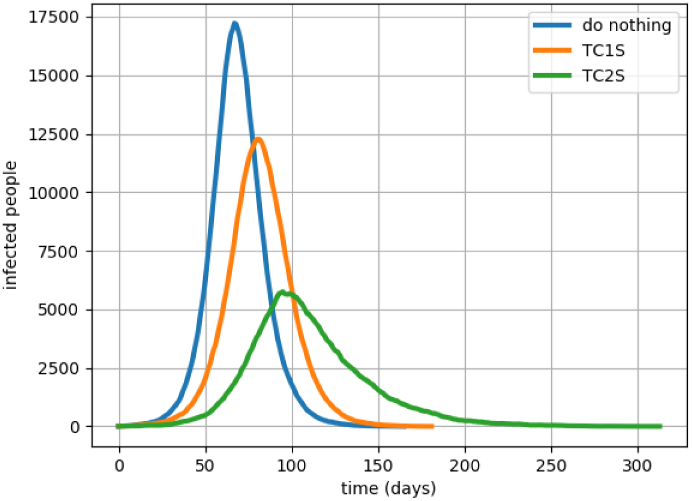
Tracing and Sealing helps but the peaks are still substantial. TC1S (trace and seal immediate contacts) does worse than the two level tracing, TC2S.

As we will see, **TC2S** works even better when it augments another strategy.

**The effect of** *p*_0_ **on the graph:** Figure 5 shows rewiring is a sensitive parameter. A moderate number of edges rewired results in speeding up the spread of the epidemic. The graphs show how fast an epidemic can sweep through the different wards of the city by simply tuning the rewiring parameter, *p*_0_.

**Fig. 5.**
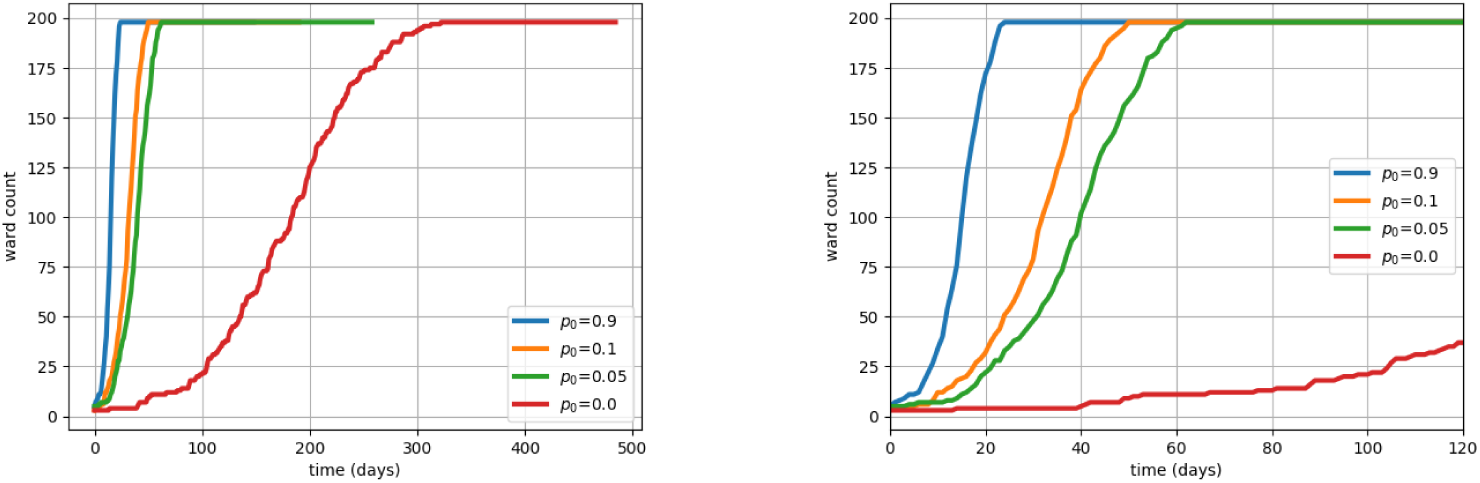
The rewiring parameter p_0_ has a role to play in how far the disease spreads across the wards. Right: Zoomed version of the graph to show even a small amount of rewiring has a huge effect on the speed of disease spread.

**The effect of relaxation factor**, *η*: The aim of most Intervention strategies is to reduce the *β*’s so that the effective transmission comes down. The relaxation factor, η, takes values between 1 and 0 and serves to reduce the transmission parameters (η*β*_M_, η*β_S_*, η*β_C_*). This allows to model various Interventions designed to hinder the transmission by a single parameter η. Figure 6 shows plots of the number of infected persons with time for various values of η. As η decreases the speed of progression becomes slower and the disease takes longer time and the peak infection also comes down. However, it effects most of the population, almost the same death and recovery rates. As reported elsewhere (for e.g. [5]), such reductions in transmission rates could be obtained by various social distancing measures. These interventions ensures that the peak infection rate comes down but overall a large fraction of population gets infected. These kind of measures help in *flattening the curve* and does not help in Suppression. The results show that the strategies of suppression belong to the regime of η < 0.5. In this regime both *R_∞_* is extremely low and length of the epidemic is reduced to below the base case. The figure in the right hand side of Figure 6 shows that the longevity of the epidemic reduces as η decreases. Thus suppression strategies should aim for Interventions which can halve the transmission rate. Such strategies would be difficult to achieve without extreme social distancing, such as Lockdowns.

**Fig. 6.**
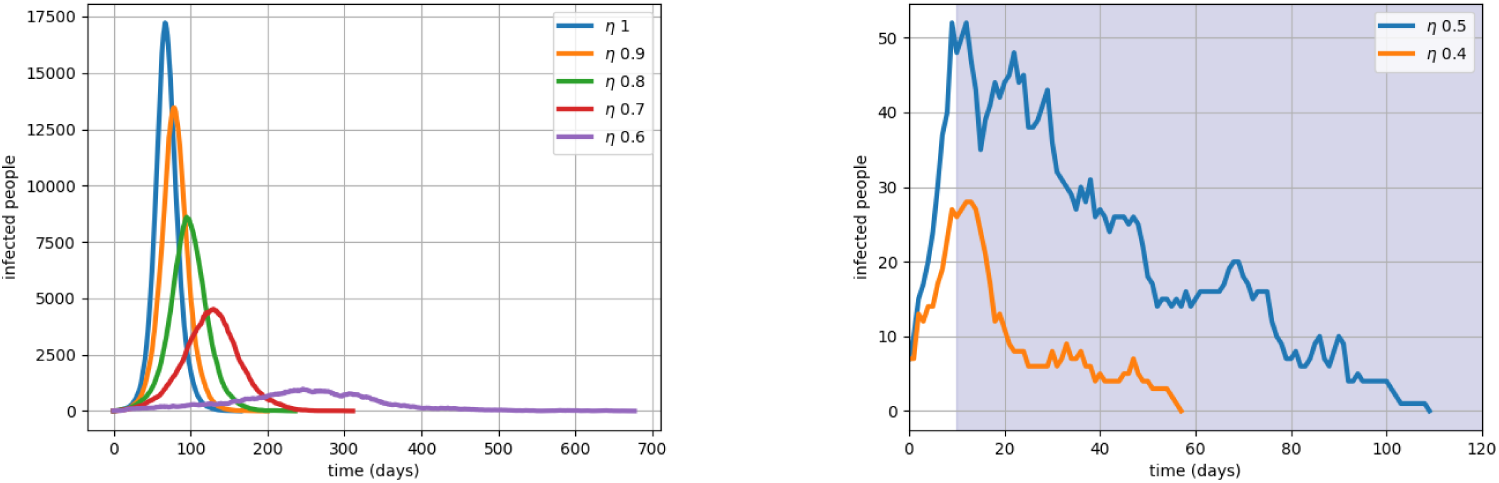
The effect of *η:* Figure on the left shows infection over time for η ≥ 0.6. Any policy which yields 1 ≥ η ≥ 0.6 will help in flattening the curve. Figure on the right shows Lockdown implemented after 20 days from the onset of the pandemic for *β* = 0.5 and *β* = 0.4. It shows that the suppression is achieved. Smaller values of η yield quicker suppression.

**Lockdowns: LD** is an extreme social distancing measure which can reduce the transmission rate. In absence of any known reliable data, we assume that a Lockdown can reduce the transmission rate to less than half. In this setup this corresponds to assuming that **LD** is a measure corresponding to η < 0.5. In particular we will use η = 0.4. Figure on the right in Figure 6 shows that it is possible that the disease dies out within 4 months. Simulation results suggest that this can be achieved by affecting only 0.52% of the population and the pandemic can end in 115 days, both values are median. Adding **TC2S** to **LD** significantly improves the infection rate by bringing it down to half. The effectiveness of any other policy can be evaluated against these set of numbers.

**Mild Lockdown:** Instead of **LD**, a mild lockdown with parameter η = 0.6 reduces *R*_∞_ to 25.75%, much lower than **DN**, but still it is much higher than that achieved by **LD**. However, if we use it along with **TC2S** then it becomes a very effective strategy, reducing the infection rate to 1%.

**Fixed duration Lockdown:** Figure 3 compares it with **DN**. Instead of seeing the peak of infection after 67 days in **DN** the new peak is shifted by 30 days approximately. It does not yield any reduction in Infection rates or in number of deaths. If the policy is to be used only once ththen it should not be used early. To avoid the peak of infection one should apply the **FDLD** a little later. The exact time to apply such a policy is still open. [3] recommends that it should be used closer to the peak. This will avoid the second peak but as a policy **FDLD**is not acceptable as it will anyway lead to significant infections and many deaths.

**Two day work week with 5 day Lockdown:** Figure 8 shows **TDWW** policy is better than mild lockdown. This is not surprising as the lockdown is withdrawn only for 2 days but is enforced for the remaining 5 days. It is still not practical as it gives a very high value of *T_epi_* and also a infection rate of more than 12%. However when enforced with **TC2S** it becomes an acceptable policy with *R*_∞_ coming down to 0.35 and it takes the same time as **LD**.

**Ward level Interventions: WSO**(0,3) policy perfoms extremely well when strict lockdowns are enforced inside the wards. The policy lowers the infection rate to 2/3 of that achieved by **LD** with similar values of *T_epi_*. As soon as the infection rises the respective affected wards are locked down. Figure 7 shows a typical run of **WSO**(0,3). It exhibits multiple peaks of infection but the number of infections at the peak is much lower than **DN**. The *W_l_* loss value is 16.52% implying that most of the wards are open most of the time. Coupling with **TC2S** further lowers the infection rate by a factor of 3 and also halves *T_epi_*. As an icing on the cake it reduces the W_l_ loss value by almost a factor of 3. However, if the lockdowns are not enforced strictly the policy can be dangerous. It is slightly better than **TC2S** and moreover with a W_l_ value of 3.77 it can be interpreted that policy results in closure of the large parts of the city for a prolonged period of time. The mild lockdown version of **WSO**(0,3) is significantly improved if used alongside with **TC2S**.

**Fig. 7.**
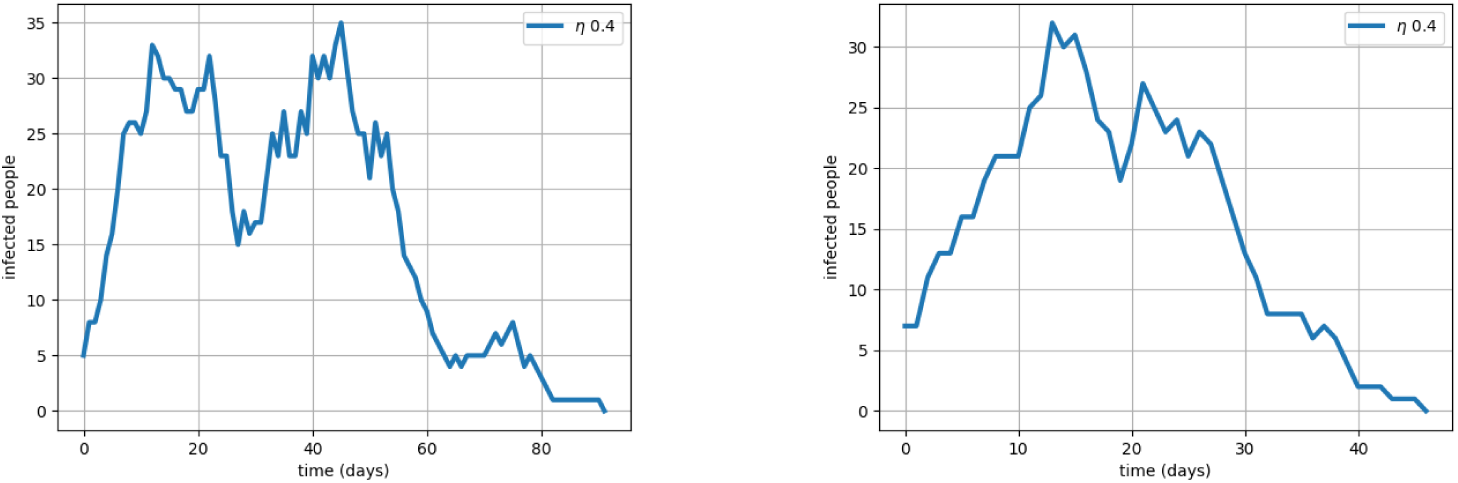
**WSO**(0,3): On the left, different wards lockdown at different times and these account for multiple peaks. On the right, when **TC2S** is imposed as well.

**Fig. 8.**
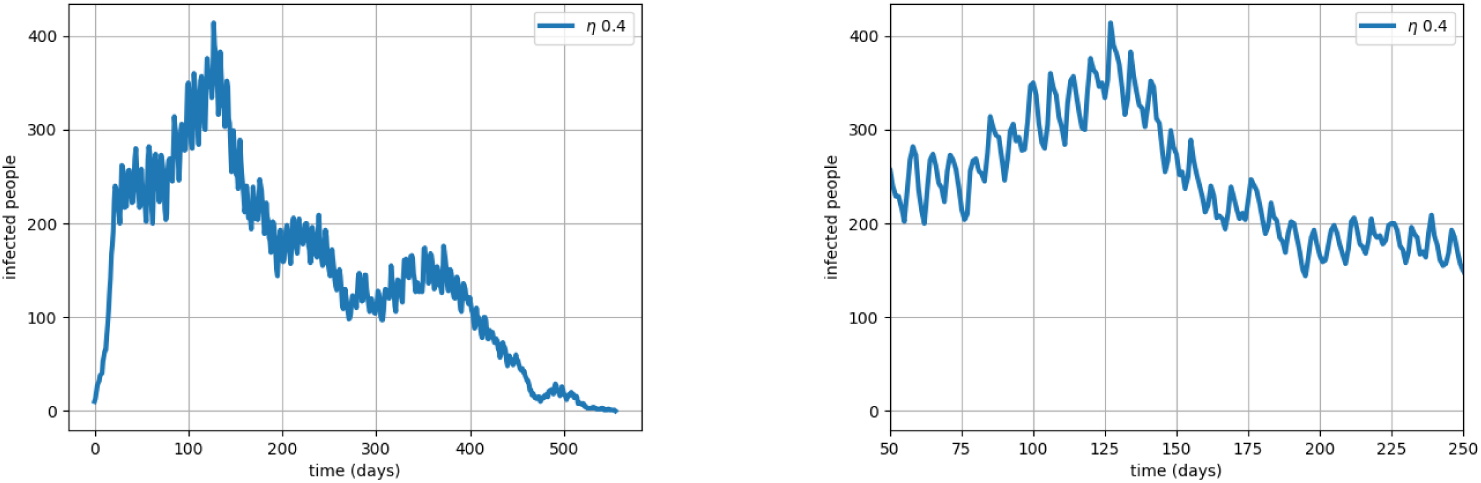
**TDWW**-policy: it prolongs the epidemic. Also, on the right is the zoomed version where one can clearly see oscillations due to lifting and imposing of ban.

## DISCUSSION

Important insights drawn from the study are summarized by the following points.

1. Doing Nothing (**DN**) and allowing infection to spread in the hope of growing *Herd Immunity* is clearly not acceptable as it results in far too many infections and death.
2. The study confirms that Lockdown(**LD**) is an acceptable strategy with and without contact tracing as it reduced the attack rate to 0.52% compared to 85% with no interventions. When used with **TC2S** it further improves to 0.35%.
3. Lockdowns of limited duration or relaxed Lockdowns are not acceptable even in the early stages. However, Lockdowns of limited duration is extremely useful to bring down the infections temporarily.
4. Releasing the lockdown for a few days in a week(**TDWW**) can be considered but it can be risky. Contact Tracing should be a must for such policies.
5. **TC2S** when used as a stand-alone strategy may not be effective but when used with some of the other alternatives it can generate much improved policies. Mild lockdowns, which on its own is not acceptable, but when coupled with **TC2S** produces an acceptable policy. Another intervention, shortened week with lockdown (**TDWW+LD**) is risky but when applied with **TC2S** becomes very effective. When used with **TC2S** it reduces the attack rate by 1/3 that of Lockdown. The effectiveness of **TC2S**, as proposed here, would depend on not only identifying the complete list of infected people in the contact network but also would require identifying any infected people in the network of the contacts. These findings are similar to [9,11] where it is argued that extensive contact tracing and case isolation may help
6. **WSO**(0,3) appears to be the most promising alternative to **LD**. It not only outperforms **LD** on attack rate but also reduces the duration of the epidemic by 1/2 when augmented with contact tracing. Even mild lockdowns with contact tracing in **WSO** can be an acceptable alternative.

## PREFERRED SUPPRESSION STRATEGY ALTERNATIVES TO LOCKDOWN

Figure 9 shows a bar-chart of preference scores, score(**A**; **LD**) (defined in (3)) of all considered policies. Any policy with score less than 1 should be preferred over **LD**. From the chart two distinct policy alternatives emerge which could match the effectiveness of Lockdown when applied as Intervention measures in early stages of the pandemic.

1. Shortened work week (**TDWW**+**LD**+**TC2S**) can be considered as an alternative to full Lockdown. The policy advocates a 2 day week where the economy is open but the remaining 5 days a week, a lockdown is enforced.
2. Opening and closing hotspots depending on Infection levels (**WSO**) can be a better alternative to **TDWW**. This policy only constrains the residents inside the ward and rest the remaining populance go about their business. **TDWW** would enforce lockdowns for most of the week and hence it is more constraining than **WSO**. Our study shows that if implemented in early stages very few wards need to be shut down.

**Fig. 9.**
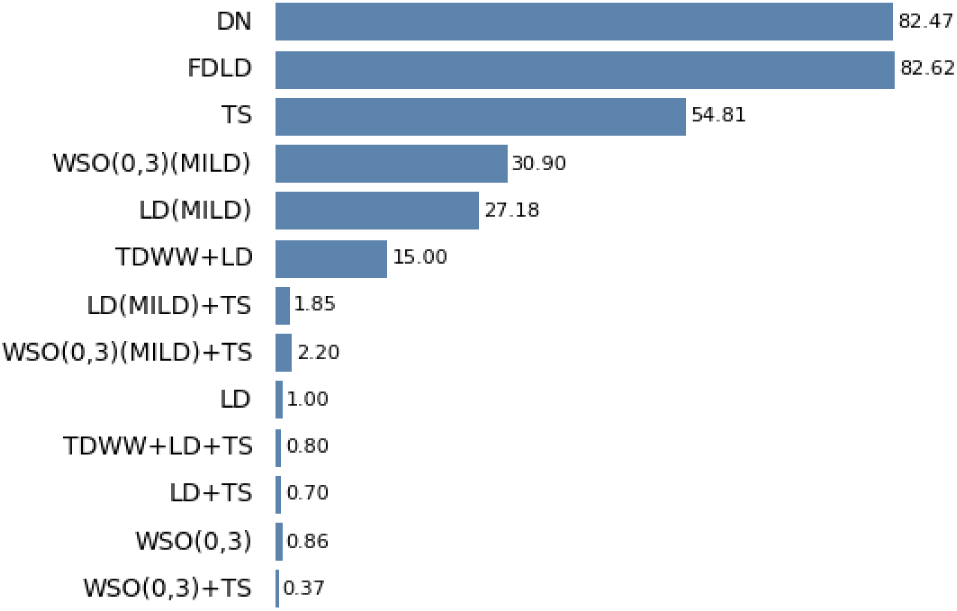
Interventions and their preference scores, score(**A**; **LD**). Smaller score indicate that policy A is better than **LD**.

The success of **WSO** policy crucially depends on the ability to Test all residents in the hotspots. While Contact tracing is not necessary it will definitely help in improving the **WSO** policy outcomes. An interesting insight is since we can target the tracing to the ward it even requires less number of contacts to be traced then implementing **LD**. If there is substantial shortage of Testing kits making **WSO** unviable then one can consider the alternative policy of opening the economy for Two days a week. If the two policies are weighed in terms of their economics then one can say that **WSO** though opens the economy much more than the 2 day week policy but also requires some investment in Testing, which could be a challenge in densely populated urban wards of metropolises. But for both the interventions require Contact Tracing.

## CONCLUSIONS

In this note we reported an empirical study of various policy alternatives to Lockdown for *suppressing* COVID19 pandemic on a city modeled by **MLSW**, a small world model. This type of small world model does not seem to have been investigated before and a detailed mathematical study of the properties should be an important area of investigation. Our results indicate three interesting highlights. Firstly, *Tracing the Contacts and their Contacts* shows substantial benefits over the usual practice of *Tracing the Contacts*. Secondly, opening the economy partially, say 2 days a week followed by a Lockdown, could be more effective than Lockdown. Thirdly, and most importantly, the most preferred strategy should be to do hotpot surveillance. It is argued that not only it has lower attack rates, it also is much faster in destroying the virus. On top of these two things it is also economically far more efficient. If this is introduced early enough it is possible that the pandemic can be suppressed in the early stages at a far lesser time then the currently practiced Lockdowns.

## Data Availability

This is a simulation model built from publicly available data. In the paper we explicitly state all the relevant data used.

## ACKNOWLEDGMENTS

We thank Prof. Rajesh Sundersan and Mr. Preetam Patil for discussion and sharing Data about Bengaluru.

## Footnotes

1 http://www.euro.who.int/en/health-topics/health-emergencies/coronavirus-covid-19/news/news/2020/3/who-announces-covid-19-outbreak-a-pandemic

